# Plasma proteomics in the prediagnostic phase of Parkinson’s disease: a multi-cohort study of 74,000 participants

**DOI:** 10.1101/2025.07.30.25332433

**Authors:** Jan Homann, Alexander G. Smith, Sarah Morgan, Elisabet A. Frick, Fangyu Liu, Vivian Viallon, Rashmi Maurya, Roxanna Korologou-Linden, Valerija Dobricic, Olena Ohlei, Laura Deecke, Fatema Hajizadah, Yujia Zhao, Fanny Artaud, Karl Smith-Byrne, P. Martijn Kolijn, Jose Maria Huerta, Nils Winter, Marcela Guevara, Ana Jimenez-Zabala, María José Sánchez, Camino Trobajo-Sanmartín, Natalia Cabrera-Castro, Ana Vinagre, Dafina Petrova, Sabina Sieri, Tim J. Key, Nick Wareham, Rudolph Kaaks, Ruth C. Travis, Tim Hahn, Susan Baker, Sean M. Kelly, Roel Vermeulen, Susan Peters, Giovanna Masala, Carlotta Sacerdote, Nancy Finkel, The Global Neurodegeneration Proteomics Consortium (GNPC), Adam S Butterworth, Alexis Elbaz, Moritz Hess, Verena Katzke, Lars Bertram, Vilmundur Gudnason, Oliver Robinson, Honglei Chen, Lefkos Middleton, Laura M. Winchester, Ioanna Tzoulaki, Valborg Gudmundsdottir, Keenan A. Walker, Pietro Ferrari, Elio Riboli, Marc J. Gunter, Christina M. Lill

**Author notes:** These authors contributed equally as shared second authors (Alexander G. Smith, Sarah Morgan, Elisabet A. Frick, Fangyu Liu, Vivian Viallon). For the index GNPC author list, see Supplementary Information. **Corresponding Author:** Dr. Christina Lill, MD, MSc, Translational Epidemiology Unit, Institute of Epidemiology and Social Medicine, University of Münster, Domagkstr. 3, 48149 Münster, Germany, and.

## Abstract

**Background:** Parkinson’s disease (PD) remains incurable, with a long prediagnostic phase currently undetectable by existing methods. Identifying individuals at high risk of PD would enhance our understanding of the underlying pathophysiology and will open up new preventive or early therapeutic avenues.

**Methods:** In the largest proteomic study in neurodegenerative diseases to date, we analyzed blood samples from ∼74,000 individuals across discovery and validation cohorts. In the discovery phase, we leveraged data from the European Prospective Investigation into Cancer and Nutrition (EPIC) cohort with up to 28 years of follow-up to identify incident PD cases across five European countries. Prediagnostic plasma samples from initially healthy participants underwent high-throughput proteomic profiling (7,285 protein markers) using the SomaScan platform. Cox proportional hazards models based on 4,538 participants (including 574 incident PD cases) were used to identify protein biomarkers associated with a future PD diagnosis. In the validation phase, we tested three cohorts with incident PD cases (AGES, ARIC, UK Biobank, (n=64,856; 1,034 incident cases), a case-control dataset with prevalent cases (GNPC, n=2,592), and the longitudinal Tracking PD cohort with data on PD progression (n=794).

**Findings:** In the EPIC4PD discovery case-cohort, 17 proteins that predict PD up to 28 years before diagnosis were detected (FDR=0.05). Additional proteins revealed sex-specific and time-varying effects, capturing disease progression before symptom onset. Replication in three prospective cohorts confirmed at least 12 key prediagnostic biomarkers with strong evidence, including TPPP2, HPGDS, ALPL, MFAP5, OGFR, ACAD8, TCL1A, GPC4, GSTA3, LCN2, KRAS, and GJA1. Furthermore, in the longitudinal Tracking PD cohort, HPGDS and MFAP5 also predicted cognitive decline. Notably, several of the identified PD biomarkers overlapped with those for incident Alzheimer’s disease and amyotrophic lateral sclerosis, indicating partially shared molecular signatures. A machine learning-derived PD protein risk score improved PD but not Alzheimer’s disease risk prediction in the independent AGES cohort.

**Conclusion:** Our extensive proteomics effort examining PD from the prediagnostic to the progression phase identified novel, actionable biomarkers opening new avenues for early PD risk stratification and precision medicine.

**Funding:** Michael J Fox Foundation, Cure Alzheimer’s Fund, Clinical Research in ALS and Related Disorders for Therapeutic Development (CreATe) Consortium, Michael J Fox Foundation, Interdisciplinary Centre for Clinical Research, University Münster

## INTRODUCTION

Parkinson’s disease (PD) is the most common movement disorder and the second most common neurodegenerative disease after Alzheimer’s disease (AD)^1^. It is characterized by progressive, disabling motor and non-motor symptoms, substantially reducing quality of life and life expectancy^2,3^. By the time PD is clinically diagnosed, significant neuronal loss has already occurred, and available treatments are only symptomatic, with diminishing benefits over time^1^. Consequently, the disease poses a significant burden not only on patients but also on their families and caregivers. Furthermore, its prevalence is projected to rise sharply, particularly in aging societies, leading to increasing financial and social costs^2^.

Importantly, non-specific symptoms may begin up to 20 years before diagnosis, suggesting that PD pathophysiology starts even earlier^4^. This time course points to a critical window for preventive or early therapeutic interventions that could delay or halt disease onset. However, identifying at-risk but clinically unaffected individuals remains a significant challenge. Developing a reliable PD risk score - such as one based on molecular biomarkers - will be essential to address this gap. Such a score would be especially valuable for risk stratification in vulnerable populations, including workers in rural areas with pesticide exposure or individuals with a family history of PD. By identifying those at highest risk, targeted monitoring and the initiation of early clinical and preventive trials could be implemented more effectively. In this context, high-throughput proteomic platforms such as SomaScan® (SomaLogic, Inc.) and Olink® (Olink Proteomics AB) offer new exciting opportunities to identify novel biological signatures of PD development. These methods can quantify thousands of proteins simultaneously and hold strong potential for identifying biomarkers that can detect diseases in their earliest or even preclinical stages^5–7^. In neurodegenerative diseases interest in blood-based biomarkers has grown, owing to their minimally invasive collection, easier clinical integration, and strong performance of candidate blood biomarkers in AD^8^. In PD, such markers could enhance early diagnosis, enable risk stratification in clinical trials, and support preventive strategies. However, given PD’s comparatively low prevalence (2-3% in individuals aged 65+)^1^, prospective cohorts with long-term follow-up are rare. Even more rare are cohorts with baseline blood samples - collected before PD onset - which are required to identify predictive biomarkers. This is why most prior biomarker studies have compared prevalent PD patients to controls, risking reverse causation and limiting predictive value^9^.

To our knowledge, only one prospective cohort, i.e., the UK Biobank (UKB), has hitherto been used to discover predictive PD biomarkers. Based on this resource, a recent study analyzed 2,920 plasma proteins measured with Olink technology in up to 859 incident PD patients^10^. Although 38 candidate biomarkers were identified, the findings of that study remain unvalidated in independent prospective cohorts. In addition to closing this gap, our study represents a major advance by nearly tripling the number of protein markers (n=7,285; SomaScan 7K array) analyzed in prediagnostic blood samples from ∼4,600 participants in a large, well-characterized case-cohort (‘European Investigation into Cancer and Nutrition for Parkinson’s disease’, EPIC4PD)^11^ within the EPIC study^12,13^. Overall, EPIC4PD features up to 28 years of follow-up time and 574 specialist-confirmed incident PD diagnoses. Crucially, we externally validated our top biomarkers in ∼65,000 individuals, including >1,000 incident PD cases, across three independent prospective cohorts. Overall, this resulted in the largest and most comprehensive investigation of predictive biomarkers for PD to date, both in terms of sample size and proteomic resolution. To assess these markers across the disease timeline - including diagnosis and progression - we also incorporated independent SomaScan 7K data from the Global Neurodegenerative Proteomics Consortium (GNPC; ∼300 prevalent cases and ∼2,300 controls)^14^ and the Tracking PD cohort (∼800 cases with progression data). Using the top results from these analysis arms, we also performed enrichment and network analyses to gain insights into biological pathways, and evaluated PD biomarker profiles in AD and amyotrophic lateral sclerosis (ALS) to assess their potential as shared markers of neurodegeneration (**Figure 1**). Finally, using machine-leaning we constructed and externally validated a PD protein risk score (PPRS) in EPIC4PD. This score improved risk prediction decades before clinical PD onset. Combined with traditional risk factors and markers from other molecular domains, the biomarkers identified in this study have the potential to substantially advance ultra-early PD risk stratification and diagnosis paving the way for early treatment and prevention.

**Figure 1.**
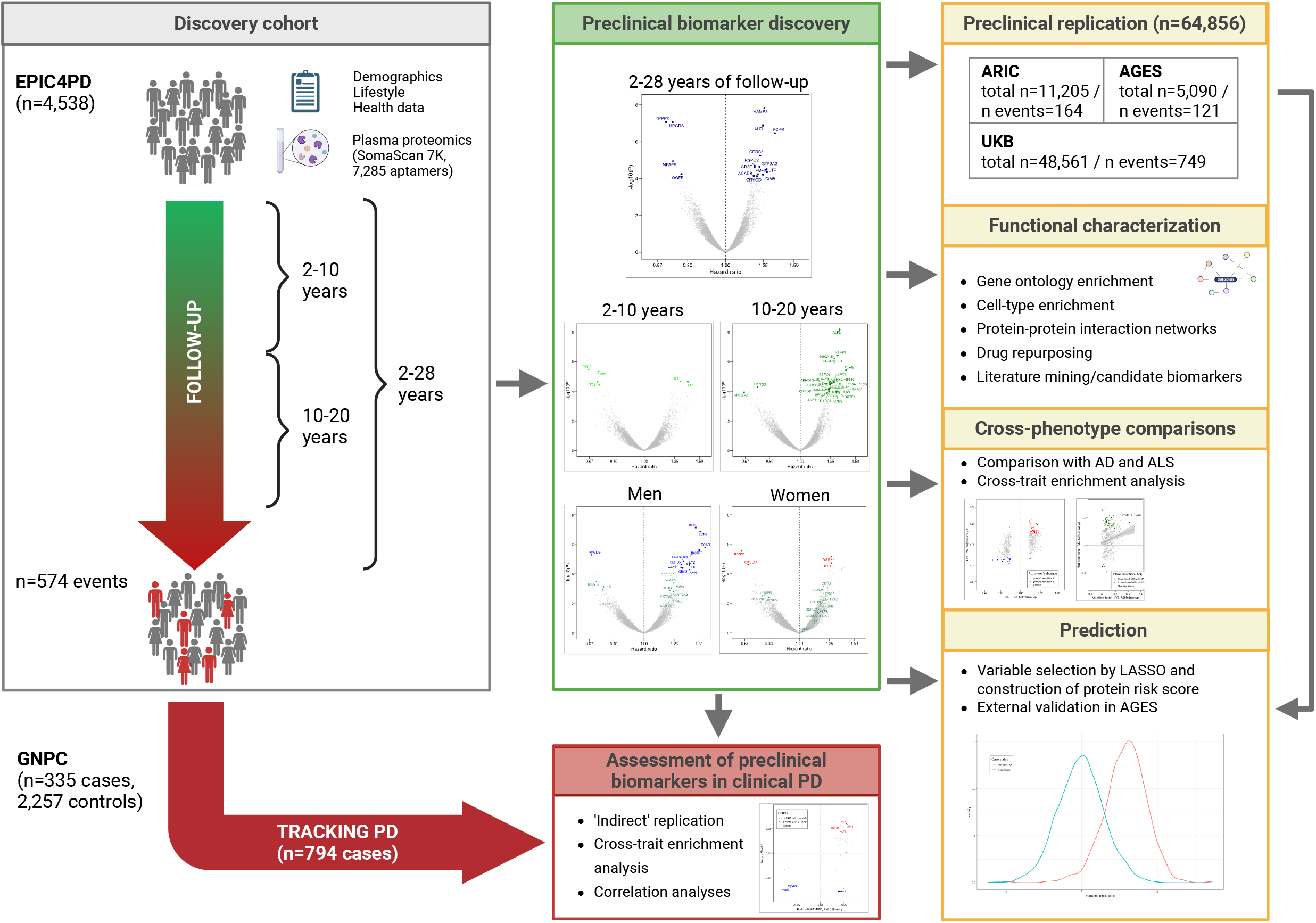
Visual summary of study design. <u>Legend.</u> This study leveraged plasma proteomics from 4,538 individuals in the ‘European Prospective Investigation into Cancer and Nutrition for Parkinson’s disease’ case-cohort (EPIC4PD) to discover prediagnostic protein biomarkers of Parkinson’s disease (PD). Using the SomaScan 7K platform, 7,285 aptamers were analyzed across multiple time windows. Identified biomarkers were externally validated in three large cohorts (‘Age, Gene/Environment Susceptibility–Reykjavik Study’ [AGES], ‘Atherosclerosis Risk in Communities’ study [ARIC], and ‘United Kingdom Biobank’ [UKB]) and assessed in clinical PD cases from the ‘Global Neurodegenerative Proteomics Cohort’ [GNPC] and the longitudinal ‘Tracking Parkinson’s disease’ cohort. Functional characterization included pathway and cell-type enrichment, protein-protein networks, cross-trait analyses with Alzheimer’s disease (AD) and amyotrophic lateral sclerosis (ALS). Predictive protein risk scores were calculated in EPIC4PD and externally validated in AGES.

## RESULTS

### EPIC4PD case-cohort

In the discovery phase of this study (**Figure 1**), we leveraged data from the EPIC cohort to build a large case-cohort (EPIC4PD) of 4,538 participants (including 574 incident PD cases; **Table 1**) across 10 European centers in Spain, Italy, the Netherlands, the UK, and Germany, combining deep proteomic profiling (6,382 proteins via 7,285 aptamers) in baseline, i.e, pre-disease plasma samples, with long-term prospective follow-up. EPIC4PD participants had a median baseline age of 53 years (range: 35–79), comprising 63% women (57% excluding the Utrecht center, which recruited only women), and were followed for up to 28 years (median: 16 years). Among PD cases, the median time from blood sampling to diagnosis was 11 years (range: 2-25), enabling biomarker discovery long before clinical onset. The age distribution at diagnosis in women and men was highly similar (median diagnosis age in each group: 71, interquartile range 65-76). As expected and consistent with known PD risk profiles, PD cases were older at baseline (median: 59 vs. 52 years), more likely male (58% vs. 40%, excluding Utrecht), less likely to be current smokers (15% vs. 25%), and slightly more often overweight (64% vs. 60% with body mass index (BMI) ≥25) compared to non-cases (**Supplementary Tables 1–2**).

**Table 1.** Overview of cohort datasets included in this study.

| Cohort | Population | Phenotype | Platform | Cases |  |  | Non-cases |  | N total |
| --- | --- | --- | --- | --- | --- | --- | --- | --- | --- |
|  |  |  |  | Number (%<br>women) | AAE (range) | AAD (range) | Number (%<br>women) | AAE (range) |  |
| EPIC4PD | Europe | incident PD | SomaScan 7K | 574 (45%) | 59 (38-77) | 71 (45-89) | 3,964 (66%) | 52 (35-79) | **4,538** |
| ARIC | USA | incident PD | SomaScan 5K | 164 (41%) | 59 (48-68) | 75 (56-87) | 11,041 (56%) | 57 (46-70) | **11,205** |
| AGES | Iceland | incident PD | SomaScan 7K | 121 (38%) | 78 (69-88) | 84 (72-93) | 4,969 (58%) | 78 (68-98) | **5,090** |
| GNPC | Europe, USA | prevalent PD | SomaScan 7K | 335 (37%) | 75 (49-90) | - | 2,257 (60%) | 71.5 (27-90) | **2,592** |
| TRACKING PD | UK | PD progression | SomaScan 7K | 794 (34%) | 66 (38-85)* | 66 (38-85) | - | - | **794** |
| EPIC4AD | Europe | incident AD | SomaScan 7K | 652 (71%) | 59 (36-69) | 78 (53-93) | 1,274 (63%) | 49 (35-69) | **1,926** |
| EPIC4ALS | Europe | incident ALS | SomaScan 7K | 187 (57%) | 58 (38-76) | 73 (48-95) | 4,528 (66%) | 52 (35-79) | **4,715** |
| UK Biobank | UK | incident PD | Olink Explore 3072 | 749 (37%) | 64 (41-70) | 71 (44-83) | 47,812 (54%) | 58 (40-70) | **48,561** |
| **TOTAL** |  |  |  | **3,572** |  |  | **70,523** |  | **74,095** |

Legend. This table presents the cohort datasets included in the study. AAE = median age at examination (baseline), AAD = median age at diagnosis. Note that the individual EPIC-derived case- cohorts (EPIC4PD, EPIC4AD, EPIC4ALS) the vast majority of non-cases, thus the total number of non-cases is not the sum of non-cases of the individual datasets. See Methods for full cohort name abbreviations. \*Mean age at visit 1.

### Association analyses in the full EPIC4PD dataset

We used Cox proportional hazards regression models across 7,285 aptamers to identify proteins associated with a future PD onset (**Supplementary Table 3**). In the basic model (stratified by age in 5-year intervals, sex, and center) covering the full follow-up period (2–28 years), 17 proteins were significantly associated with PD risk after a false-discovery rate (FDR) control of 0.05 (**Table 2, Supplementary Table 4**). We applied two approaches in the discovery phase to address outliers: one excluded extreme values as potential noise, and the other retained them via capping, as they may represent strong PD signals. Nine proteins were FDR-significant in both approaches; for the other eight (six upon exclusion, two upon capping), the alternative model still showed consistent, nominally significant associations with smiliar effect size estimates (mean ΔHR⍰=⍰0.03, maximum ΔHR=0.13; **Supplementary Figure 1A**), supporting robustness of our findings. For simplicity, we report only one result (from the better-performing model) in the main text, with capped data labeled “cap”; full results are available in **Supplementary Table 3**. Top proteins associated with PD incidence included VAMP3 (hazard ratio [per 1 standard deviation (SD) increase in log_10_-transformed protein levels]=1.26, p=1.46E-08), TPPP2 (HR=0.70, p=8.52E-08), HPGDS (HR=0.73, p=8.59E-08), ALPL_cap_ (HR=1.25, p=1.30E-07), FCAR (HR=1.34, p=3.48E-07), and CERS5 (HR=1.23, p=5.64E-06; **Figure 2A+D, Table 2**). All FDR-significant associations remained stable after additional adjustment for key demographic and lifestyle factors (education level, BMI, smoking, and physical activity), with minimal impact on effect sizes (mean ΔHR=0.02, maximum ΔHR=0.07; **Supplementary Figure 1, Supplementary Table 4**).

**Table 2.**
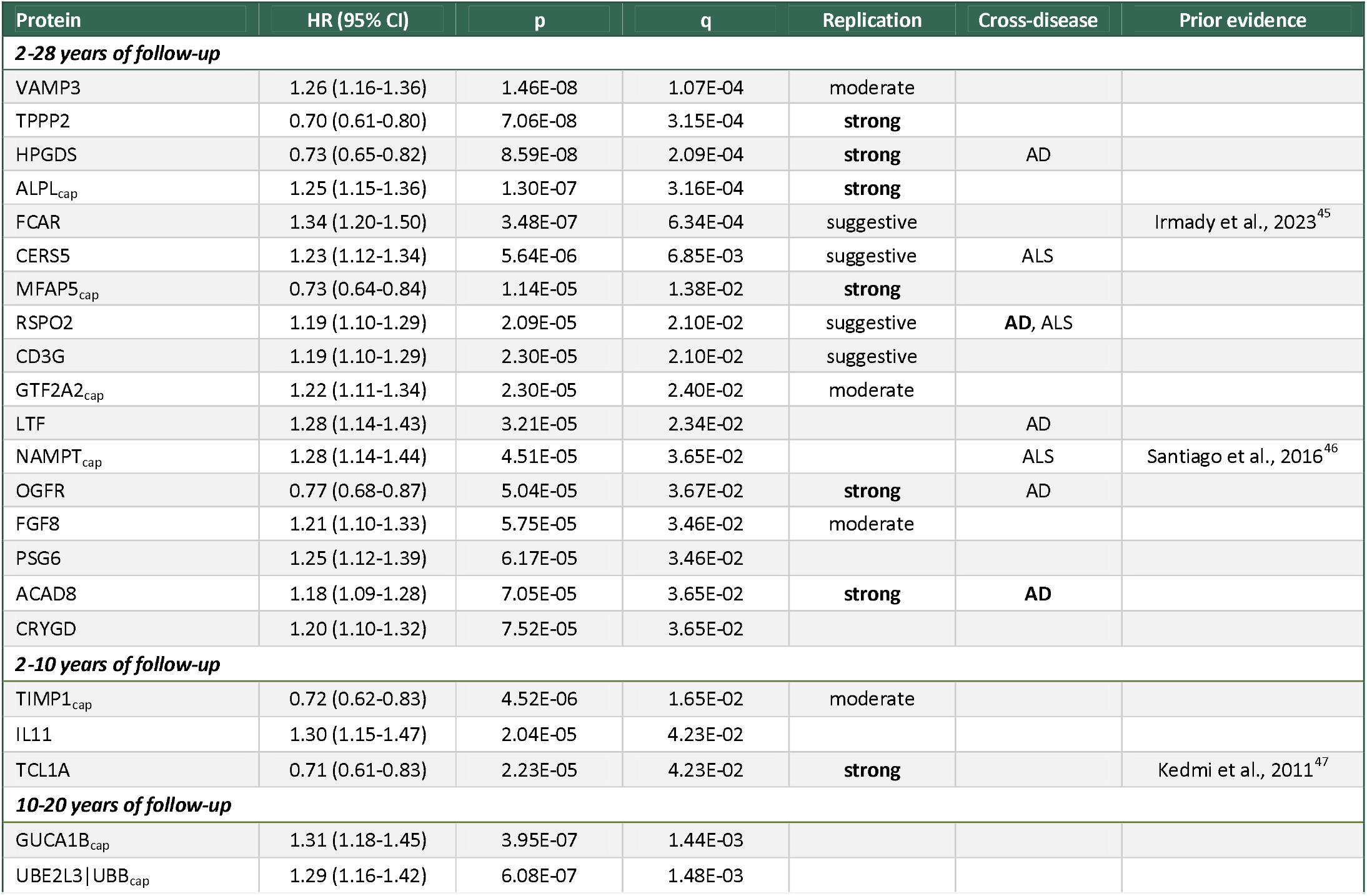
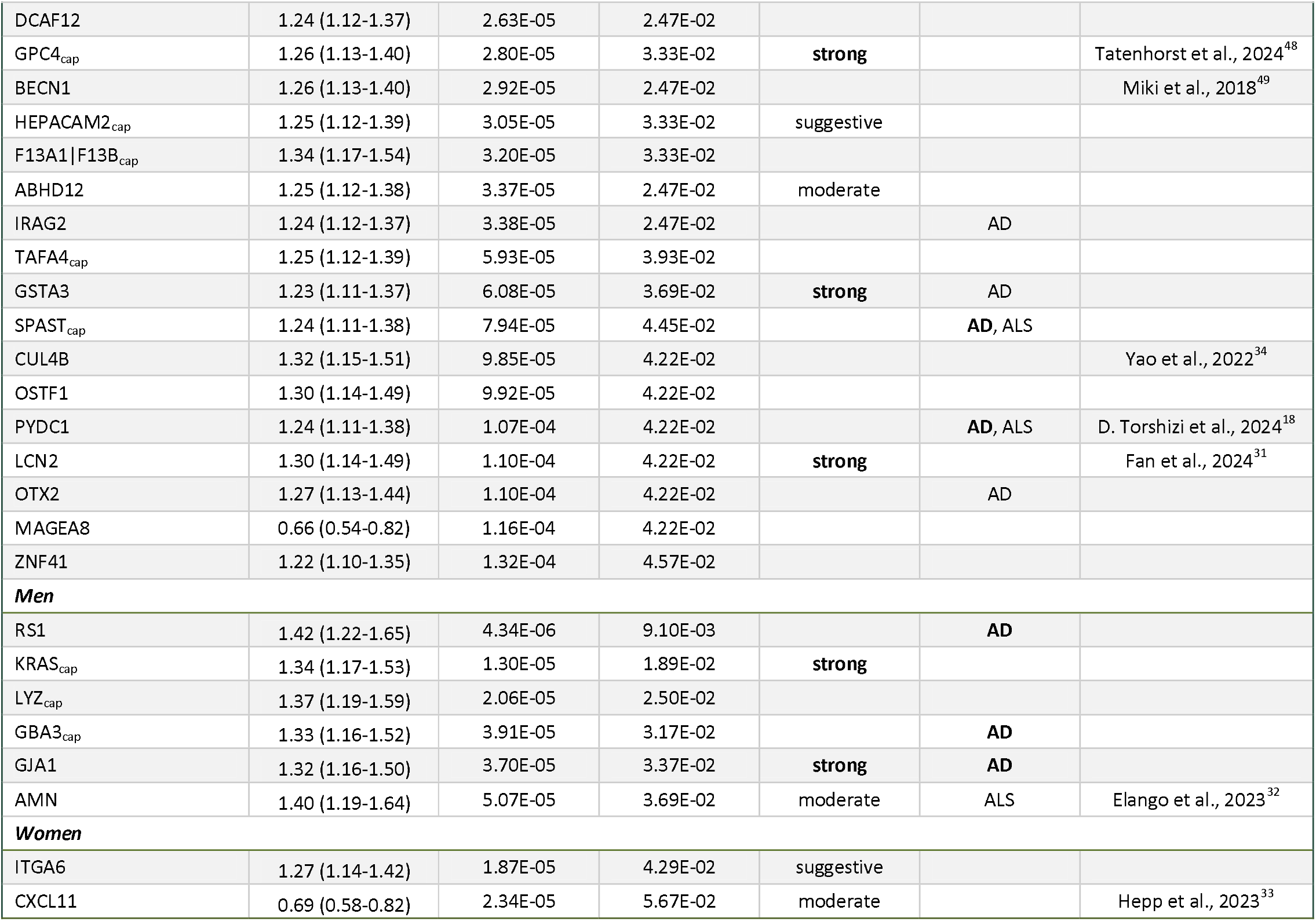
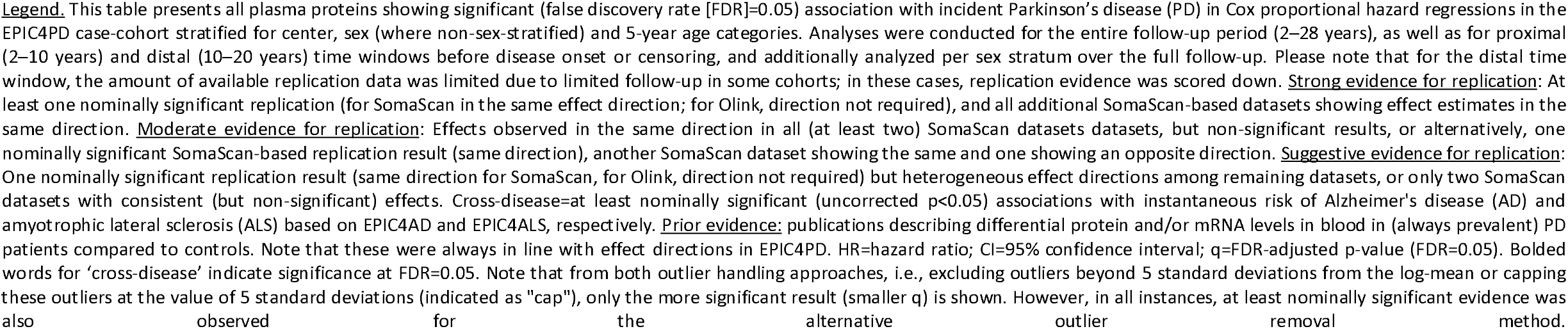
Plasma protein biomarkers significantly associated with future onset of Parkinson’s disease.

**Figure 2.** Cox proportional hazard regression analyses across different time-windows in EPIC4PD. <u>Legend.</u> Association results of prediagnostic protein biomarkers with incident Parkinson’s disease across different follow-up windows in EPIC4PD. Panels **A–C:** effect estimates for three time windows: full follow-up (2–28 years), proximal (2–10 years), and distal (10–20 years). **(D)** summary of associations of significant biomarkers (false-discovery rate of 0.05 in at least one time-window) across all follow-up periods.

### External replication of predictive biomarkers in the full follow-up period

Of the 17 FDR-significant biomarkers identified in EPIC4PD, replication data were available in a total of 64,856 participants (including 1,034 incident PD cases) from three prospective studies: the Gene/Environment Susceptibility–Reykjavik Study (AGES), Atherosclerosis Risk in Communities study (ARIC), and UKB. Specifically, all 17 proteins were available in AGES (5,090 participants, 121 cases; SomaScan 7K), 15 in ARIC (11,205 participants, 164 cases; SomaScan 5K), and 8 in UKB (48,561 participants, up to 749 cases; Olink 3072 Explore; **Supplementary Table 5, Supplementary Figure 2**). The other 9 proteins were not measured in UKB, as the current Olink platform captures only ∼30% of the proteins assayed by SomaScan 7K. We also included independent SomaScan 7K-based data from prevalent cases as ‘indirect’ replication. To this end, we re-analyzed proteomic profiles from 2,592 participants (335 prevalent PD cases and 2,257 controls) across three GNPC case-control datasets (also see below for comparison of incident and prevalent PD), using similar methodology as with our discovery phase analyses.

Given the higher comparability of results from datasets generated using the same technology, we first considered replication evidence in the SomaScan-based datasets (AGES, ARIC, and GNPC). Overall, nine of the 17 proteins - TPPP2, HPGDS, ALPL, MFAP5, OGFR, ACAD8, VAMP3, GTF2A2, and FGF8 showed strong or moderate evidence for replication, i.e., nominally significant evidence for association in at least one dataset and/or fully consistent effect estimates across all independent studies with available data (**Figure 3A, Table 2, Supplementary Table 5**). Upon reviewing the replication evidence in the UKB, the evidence of association was further substantiated for TPPP2, HPGDS, MFAP, and OGFR, while ALPL, ACAD8, VAMP3, GTF2A2, and FGF8 were not measured by Olink. Several additional proteins showed suggestive replication evidence in individual datasets, but the overall replication evidence was more heterogeneous (**Table 2, Supplementary Figure 2, Supplementary Table 5**). Overall, platform-related differences between Olink and SomaScan must be considered.

**Figure 3.** Validation and characterization of top prediagnostic biomarkers. <u>Legend.</u> **(A)** Forest plot of results of prediagnostic biomarkers for incident PD in the EPIC4PD case-cohort, the ‘Age, Gene/Environment Susceptibility–Reykjavik Study’ (AGES), the ‘Atherosclerosis Risk in Communities’ study (ARIC), and ‘United Kingdom Biobank’ [UKB] cohorts. **(B)** Comparison of top prediagnostic PD biomarkers with their associations in prevalent PD within the ‘Global Neurodegenerative Proteomics Cohort’ (GNPC); left: false-discovery rate (FDR)-significant proteins from EPIC4PD; right: nominally significant proteins. Color-coding refers to risk (red) or protective (blue) effects in GNPC. **(C)** Cross-disease analysis of prediagnostic PD-associated proteins compared to incident Alzheimer’s disease (AD) and amyotrophic lateral sclerosis (ALS) in EPIC4AD and EPIC4ALS. The top panels display all FDR-significant prediagnostic PD proteins from the full follow-up analysis, while the bottom panels show proteins with nominal significance (p<0.05). Protein names in orange indicate nominal significance in AD (top left) or ALS (top right). Orange bolded names indicate FDR-significant proteins. For each disease, the bottom right plot illustrates the correlation of effect estimates using ‘mirrored’ beta values (see Methods). **(D)** Protein-protein interaction network of FDR-significant predictive proteins (black border) and GWAS-identified hits (no border). Eight clusters were identified using igraph’s greedy optimisation of modularity. Node size increases for nodes with 10 or more edges, proportional to the number of edges. **(E)** Integrated overview of datasets for validation and characterization of FDR-significant biomarkers across different proteomic and transcriptomic layers.

In line with the previous UKB study^10^, we observed largely decreased protein signals in incident PD cases in our re-analyses of the UKB Olink data (i.e., 87% of nominally significantly associated proteins) – a pattern was not seen in SomaScan-based datasets from EPIC4PD, AGES, or ARIC (proportion of proteins with a decreased protein signal for incident PD: 43% to 56%; **Supplementary Table 6**), underscoring the challenges of comparisons across different protein platforms (see **Discussion**).

In summary, nine of the 17 biomarkers (53%; ACAD8, HPGDS, TPPP2, OGFR, MFAP5, ALPL, FGF8, VAMP3, GTF2A2) were replicated across different technologies, populations, and study designs, reinforcing their specific predictive potential, and, more generally, support the generalizability of our biomarker signals.

### Biomarker associations relative to disease onset

Stratifying analyses by temporal proximity to PD diagnosis revealed remarkable time-varying biomarker effects. In the window closest to disease onset (2-10 years; n=4,230 participants including 245 incident cases), three additional proteins - IL11, TCL1A, and TIMP1_cap_ - emerged as FDR-significant. In the more distal window (10-20 years; n=4,023 including 307 incident cases), 19 additional proteins showed strong associations with PD incidence (**Figure 2B+C, Table 2, Supplementary Table 4**). These proteins had not been identified as significant overall due to negligible effects in the other time window (**Supplementary Table 3**). Time-varying biomarker signals were further supported by HR pseudotrajectories derived from 5-year sliding windows (**Figure 2B+C, Supplementary Figure 3**). Consequently, we performed additional exploratory statistical analyses, including interaction analyses with time-to-event and spline regressions to assess non-linear association effects. Importantly, a substantial number of biomarkers (all biomarkers from the proximal window and nine biomarkers from the distal window) demonstrated nominally significant interaction results and/or evidence from spline regressions for non-linear associations, providing enhanced statistical support for time-varying effects (**Supplementary Table 7, Supplementary Figure 4**). Together, these findings demonstrate that key biomarkers for PD may emerge at distinct prediagnostic phases, underscoring the value of deep proteomic profiling with extended follow-up to reveal dynamic, stage-specific signatures of prediagnostic disease.

### External replication of the temporal analysis results

Of the three newly emerging biomarkers from the proximal follow-up window (2-10 years before diagnosis), TCL1A showed strong independent replication evidence and TIMP1 showed moderate support, while IL11 did not replicate (**Figure 3A, Supplementary Figure 2, Supplementary Table 5**). Generally, the amount of replication data for the distal time window (10-20 year window) was more restricted due to shorter follow-up durations in some of the replication cohorts **(Methods** and **Supplementary Table 5**). Among the biomarkers emerging from the 10-20 year window, GPC4, GSTA3, and LCN2 exhibited strong evidence of replication, and ABHD12 showed moderate support (**Table 2, Figure 3A**). For LCN, we observed nominally significant evidence for replication in ARIC with the same effect direction as in EPIC4PD. In UKB, LCN2 also reached nominal significance (p=3.80E-03) but showed an inverse direction of effect (**Figure 3A, Supplementary Table 5**). This discrepancy likely reflects methodological differences between the SomaScan and Olink proteomic platforms (see **Discussion**).

### Sex-stratified analyses

In EPIC4PD, 18% (3 of 17) of proteins significantly associated with PD risk in the full follow-up period showed clear sex-specific effect size differences and significant interaction with sex after FDR control. These proteins included NAMPT_cap_ (ΔHR⍰=⍰48%), FCAR (ΔHR⍰=⍰42%), and ALPL_cap_ (ΔHR⍰=⍰34%; **Supplementary Figure 1G, Supplementary Table 8**).

Furthermore, sex-stratified proteome-wide analyses uncovered six additional proteins (RS1, KRAS_cap_, LYZ_cap_, GBA3_cap_, GJA1, AMN) FDR-significantly associated with PD risk in men and two (CXCL11, ITGA6) in women, none of which showed strong associations in the opposite sex (HRs 0.92–1.10, all *p*≥0.05; **Supplementary Figure 1G, Supplementary Figure 5; Supplementary Table 4**). Interaction analyses confirmed FDR-significant sex-based risk modifications for all male-specific biomarkers except GJA1. However, the latter and CXCL11 still showed nominally significant evidence for interaction (**Supplementary Table 8**). The results of these 11 FDR-significant proteins in the sex-stratified analyses remained stable after adjusting for lifestyle variables and, in women, for menopausal status and postmenopausal hormone therapy (**Supplementary Figure 1D+E, Supplementary Table 4**). While the amount of replication data was more limited due to smaller strata in the replication cohorts, KRAS_cap_ and GJA1 demonstrated strong evidence of replication in men, while AMN (men) and CXCL11 (women) provided moderate evidence for replication (**Figure 3A**). Collectively, these findings underscore the importance of accounting for sex-specific effects in biomarker discovery, as such effects may reveal associations that remain hidden in sex-combined analyses.

### Inter-correlations of predictive biomarkers in EPIC4PD

Several FDR-significant biomarkers showed moderate inter-correlations (r=0.4-0.6) in EPIC4PD, suggesting shared regulation or involvement in the same pathway. Correlations were observed between MFAP5 and both HPGDS and TPPP2 (r=0.54 each) and between LCN2 and LTF (r=0.60). FCAR also featured in several correlation clusters, underscoring its broader connectivity (detailed in **Supplementary Figure 6** and **Supplementary Table 9**).

### Characterization of prediagnostic biomarkers in prevalent PD and PD progression

Next, we investigated whether the effects of the identified prediagnostic biomarkers extended beyond the risk phase into the symptomatic stage of PD by leveraging the SomaScan 7K proteomic data from 2,592 independent GNPC participants (335 prevalent cases and 2,257 controls; see the ref. ^14^ for full GNPC dataset description). Of all 47 prediagnostic biomarkers detectable in GNPC, 7 (15%; TPPP2, HPGDS, ALPL, NAMPT, GSTA3, KRAS, GJA1) were nominally significantly associated with prevalent PD, and 4 (TPPP2, HPGDS, NAMPT, GSTA3) remained significant after FDR control. All but NAMPT showed consistent effect directions. Overall, 38 (81%) of the 47 top proteins and 125 (86%) of the 146 nominally significant aptamers results shared between both traits showed concordant directions of effect - significantly more than expected by chance (p=1.25E-5 and p=1.64E-19, respectively; one-sided exact binomial test; **Figure 3B, Supplementary Table 10**). In serum samples from the Tracking Parkinson’s cohort (n=794), we assessed associations between FDR-significant prediagnostic biomarkers and clinical outcomes related to disease progression, using motor (Unified Parkinson’s Disease Rating Scale Part III, UPDRS-III) and cognitive (Montreal Cognitive Assessment, MoCA) scores in cross-sectional and longitudinal mixed-effects models. Two biomarkers, HPGDS (β=2.61, p=6.82E-04) and MFAP5 (β=1.31, p=1.49E-03), were FDR-significantly associated with higher MoCA scores over time, indicating that lower levels of these proteins predict both increased PD risk and faster cognitive decline (**Figure 3E; Supplementary Table 11**). Associations remained robust after adjusting for BMI and disease duration (data not shown). These results demonstrate that a considerable fraction of prediagnostic biomarkers persist in their dysregulation in prevalent PD, and a small subset is even associated with progression. Comparing incident and prevalent disease settings, such as between EPIC4PD and GNPC, is essential for determining which biomarkers are robust across the disease continuum and which are specific to particular stages. This distinction enables identification of biomarkers suitable for early detection versus those better suited for diagnosis or disease monitoring.

### Comprehensive assessment of previously described biomarker candidates

Of 291 unique proteins previously linked to PD across methodologically diverse sources^10,15– 17^, 191 proteins were assessable in the EPIC4PD data (**Supplementary Table 12**): Most notably, 38 proteins had recently been reported as putative predictive PD biomarkers in the UKB Olink study^10^, with 27 also covered in our independent dataset. Of these, 11 (41%; HPGDS, CD209, TPPP3, EPS8L2, FCRL1, BAG3, TNXB, IL13RA1, NEFL, PPY, RET) showed at least nominally significant associations in one or more follow-up intervals in EPIC4PD. HPGDS also represents a top biomarker in EPIC4PD (**Table 2**). The next two most significant proteins were CD209 (best model for follow-up of 10-20 years: HR=0.75, p=1.32E-04) and TPPP3 (best result: 2-28 years: HR=0.80, p=1.58E-03), which aligns well with TPPP2, one of the lead findings in EPIC4PD. All but one (PPY) of the at least nominally significantly associated biomarkers showed the same effect direction as in the previous study^10^ despite platform differences. Of the 127 candidate neurological biomarkers from the CNS NULISA panel (Alamar, Inc), 106 were tested in EPIC4PD; of these, UBE2L3|UBB and CCL11 represented top biomarkers in EPIC4PD (**Table 2**). Other nominally significant findings included for instance SNCA, DDC, NEFL, TREM2, GFAP, and TIMP3. Furthermore, PYDC1 nominated by a recent Mendelian randomization (MR) and colocalization study^18^ was among the top results in EPIC4PD suggesting a causative link to PD risk. Several other candidates from an additional MR^16^ or GWAS^17^ showed nominal associations in EPIC4PD (Supplementary Table 12). Conversely, 5 of the 39 FDR-significant PD biomarkers identified in our non–sex-stratified EPIC4PD analyses (CRYGD, LTF, HPGDS, TCL1A, and LCN2) were also included in the MR study based on SomaScan data^16^. Of these, two biomarkers (CRYGD and LTF) showed nominal evidence of a causal association with PD, suggesting that higher circulating levels may increase disease risk, in line with the EPIC4PD findings (**Supplementary Table 13**). The limited overlap in assessed proteins likely reflects the comparatively stringent analysis criteria applied in that study^16^.

### Cross-disease comparisons

The 47 FDR-significant PD biomarkers identified in EPIC4PD were also evaluated in incident AD (n=1,926, including 652 cases) and ALS (n=4,715, including 187 cases) using Cox regression analyses in the EPIC4AD and EPICALS case-cohorts (**Supplementary Table 1**), respectively. Sixteen (34%) prediagnostic PD biomarkers showed at least nominally significant associations with AD and/or ALS, all with concordant effect directions. Correlation plots of hazard ratios showed alignment in effect size estimates. Notably, three PD proteins - RSPO2, SPAST_cap_, PYDC1 - were FDR-significant for AD and also showed nominally significant associations with ALS (**Table 2, Figure 3C, Supplementary Table 14**).

At the proteome-wide level, 17% of proteins nominally significantly associated with PD over the full follow-up period (non-sex-stratified) also showed associations with AD and/or ALS, whereas 83% appeared to be PD-specific. Among the shared proteins, the majority (68%) overlapped exclusively with AD, 25% with ALS, and 7% were common to all three neurodegenerative diseases (**Supplementary Figure 7**). Nominally significant prediagnostic AD biomarkers were significantly enriched among nominally significant PD biomarkers (p=1.19E-03; Fisher’s exact test), while ALS biomarkers were not (p=0.107). Notably, 85 of 92 PD–AD and 33 of 39 PD–ALS shared proteins showed concordant effect directions (p=1.92E-18 and p=7.15E-06; exact binomial test; **Figure 3C**). Additionally, among proteins nominally associated with PD, effect sizes modestly correlated with AD and ALS (Pearson’s r=0.18 [95% CI: 0.11–0.25] and r=0.21 [0.14–0.28], respectively; **Figure 3C**). While these comparisons highlight a largely PD-specific biomarker signature, they also point to partially shared prediagnostic pathways.

### Functional characterization of prediagnostic PD biomarkers

Gene ontology (GO) enrichment analysis of suggestive PD biomarkers (n=154 proteins with p<1.0E-03 and consistent effect directions; see Methods) did not yield FDR-significant results but revealed 68 gene sets with strong nominal enrichment (p<1.0E-02), highlighting numerous biologically plausible and likely disease-relevant pathways based on prior evidence. Strikingly, despite the long time between blood draw and clinical diagnosis, we observed several well-established mechanisms in PD among the top ‘biological processes’ categories, including vesicle trafficking (e.g., endoplasmatic-reticulum-to-Golgi transport, vesicle docking), dopaminergic neuron differentiation, microtubule dynamics, and immune-related responses (e.g., to interleukin-6, cytokines, neutrophil-mediated immunity). Lastly, several vitamin-related processes, including response to vitamin D, also emerged, pointing to molecular pathways that were especially prominent in earlier PD research. Enrichments in ‘cellular component’ and ‘molecular function’ categories reinforced the role of vesicle-mediated activity, while Reactome pathway analyses further emphasized immune system involvement, including antimicrobial peptides (**Supplementary Tables 15**).

Expression heatmaps across 50 human bulk tissues and single brain cell types based on RNA-seq data revealed relatively high expression of several FDR-significant biomarkers in peripheral and central immune-related tissues and cells compared to other tissue types (**Supplementary Figures 8-9**). Protein-level data from the Human Protein Atlas supported this observation (**Supplementary Figure 10**). Consistent with these findings, single-cell RNA-seq–based enrichment analyses of FDR-significant and suggestive biomarkers in mice and humans supported a role for immune cells in prediagnostic PD. In mouse cortex, suggestive biomarkers showed nominally significant enrichment in microglia (**Supplementary Table 16**). Analyses across 23 representative human cell types (including immune, neuronal, intestinal, muscle, liver, and fibroblast cells) demonstrated nominally significant enrichment in fibroblasts, monocytes and Kupffer cells, with borderline significance in granulocytes and macrophages (**Supplementary Table 16**). Collectively, these single-cell RNA-seq–based findings align with our Gene Ontology and Reactome pathway results, highlighting early immune system involvement as a key feature of prediagnostic PD.

Interestingly, protein–protein interaction (PPI) analysis revealed that FDR-significant biomarkers were significantly more likely to interact with GWAS-nominated PD risk genes^17^ than expected by chance (p=1.0E-04; permutation test). Within this network, SNCA emerged as a central hub, exhibiting several direct edges with FDR-significant biomarkers (**Figure 3D, Supplementary Table 17**). Consistently, *SNCA* expression in both brain and immune cells was correlated with several of these biomarkers (**Supplementary Table 18**). In brain, strong correlations (r≥0.5) were observed with RS1, RSPO2, and OSTF1, and in immune cells, with LYZ, VAMP3, F13A1, and FCAR. In paired samples of healthy participants from the GNPC, several proteins (GTF2A2, LCN2, LYZ, TIMP1, OGFR, NAMPT, and CULB4B) exhibited moderate to strong correlations between blood and CSF (**Supplementary Table 19**). These findings provide converging evidence that the identified prediagnostic biomarkers capture early pathophysiological processes consistent with independent lines of prior research. Finally, a semi-systematic literature search revealed prior functional evidence for a major part of the 47 prediagnostic PD biomarkers (details in the **Supplementary Material** [**Supplementary Table 20**]).

### Druggability assessment of prediagnostic biomarkers

Given their potential pathophysiological implications, we next evaluated whether any of the 47 FDR-significant prediagnostic PD biomarkers represent druggable targets using the Open Targets database^19^. Twenty-one proteins (45%) showed at least one supporting line of evidence for interaction with small-molecule drugs (**Supplementary Table 21**). Notably, four proteins - NAMPT, KRAS, CD3G, and F13A1 - were associated with compounds currently undergoing clinical trials in non-neurological diseases (mostly cancer), underscoring their translational potential. These findings indicate that a substantial share of prediagnostic PD biomarkers may be pharmacologically modifiable.

### Disease prediction

To assess predictive utility, we fitted penalized Cox models based on least absolute shrinkage and selection operator (LASSO)-based machine learning in EPIC4PD, excluding the Heidelberg center as training and using it as test dataset. The external validation of the best performing PD protein risk score (PPRS) from the test data (as determined based on the concordance [C-]index) in the independent AGES cohort (n=5,090; 121 incident PD cases), which also used the SomaScan 7K platform, yielded an improvement of the C-index of 0.07, i.e., from 0.65 (baseline based on age and sex only) to 0.72 (baseline + 8 proteins) in the overall sample. This improvement is statistically highly significant (p=4.66E-09, likelihood ratio test). Three of the eight proteins from the PPRS (i.e., TPPP2, HPGDS, VAMP3) also appear as top results in the single-protein analyses (**Supplementary Tables 22-23**). In AGES, the PPRS translates to a nearly 6-fold increase of PD risk in the top compared to the bottom PPRS quintile during the AGES follow-up of up to 17 years prior to disease diagnosis (HR=5.70, p= 6.62E-07). Importantly, for incident AD in AGES, the increase in C-index using the eight-protein PPRS was negligible (ΔC-index=0.0019), suggesting specificity of the score for PD despite overall concordant effect directions across the two neurodegenerative endpoints (**Supplementary Table 23**).

## DISCUSSION

In this large, prospective, and multi-center proteomic study, we identified and independently validated novel plasma biomarkers predictive of PD onset up to 28 years before clinical diagnosis. Using the high-resolution SomaScan 7K platform in prediagnostic samples from over 4,500 EPIC4PD participants, we discovered 47 proteins significantly associated with incident PD. Several top markers - TPPP2, HPGDS, ALPL, MFAP5, OGFR, ACAD8, TCL1A, GPC4, GSTA3, LCN2, KRAS, and GJA1 - showed strong replication across incident (AGES, ARIC, UKB) and prevalent (GNPC) PD cases, underscoring their robustness across platforms and populations. Importantly, owing to the much larger proteomic resolution our study uncovered numerous novel PD biomarkers not assayed in the previous UKB analyses^10^. In addition, while we observed that prediagnostic protein biomarker profiles are partially shared between PD and other neurodegenerative diseases (i.e., AD and ALS), the delineated PD risk score appears to be disease-specific. Overall, our study delivers the most comprehensive proteomic resource for prediagnostic PD to date. It is unique by incorporating diverse prediagnostic validation cohorts, by spanning the full disease course, and by demonstrating some overlap with the prediagnostic phase of other neurodegenerative disorders. As such it sets a new benchmark for biomarker discovery in neurodegeneration.

Our findings offer several new key insights: First, predictive PD biomarkers can be reliably identified in blood many years, even decades, before diagnosis. In this context, signals such as HPGDS, MFAP5, OGFR, and ACAD8 appear to remain stable across different populations and proteomic platforms. Furthermore, we also observed dynamic, stage-dependent biomarker association patterns: while some protein signals persist throughout all stages, others appear only within certain time windows before diagnosis (such as IL11, TIM1, and TCL1A in the 2-10 year window, and several others in the 10-20 year window prior to diagnosis), possibly reflecting underlying pathophysiological shifts. Interestingly, all three biomarkers from the proximal time window are linked to immune processes and inflammation (as detailed in the **Supplementary Material**). Furthermore, it is notable that we observed a high degree of agreement (>80%) in effect size directions between prediagnostic and clinical biomarkers across the independent EPIC4PD and GNPC datasets (both SomaScan 7K). Additionally, two of the prediagnostic biomarkers (HPGDS and MFAP5) also predicted longitudinal cognitive decline in the course of PD. These comparisons - which uniquely span the broad spectrum from preclinical PD to disease progression - enable the identification of biomarkers relevant across all stages versus those that are stage-specific, thereby informing their optimal use for early detection, diagnosis, and disease monitoring. Second, we observed evidence for sex-specific effects for several biomarkers in EPIC4PD, with predominance in men, which were not driven by other relevant covariates such as smoking or BMI. This is interesting in light of the increased prevalence of PD in men and warrants detailed follow-up in future investigations.

Third, the substantial concordance of findings across different independent SomaScan-based datasets supports the robustness of our findings. In this context, we note that the previous UKB study^10^ reported a consistent decrease of top prediagnostic biomarkers in incident PD cases when compared to non-cases. While we were able to confirm this pattern in our re-analysis of the publicly available UKB data, we did not observe this in any of the four SomaScan-based datasets. Likely, this discrepancy reflects methodological differences between the currently available proteomic platforms: While recent evidence suggests that the latest SomaScan platforms provide higher measurement precision when compared to Olink^20–24^, it is important to note that both platforms frequently capture distinct protein features, underscoring their complementary strengths and limitations^20^ (see **Supplementary Text** for a more detailed discussion on this). A better understanding of these differences will require large paired SomaScan and Olink datasets as well as benchmarking against mass spectrometry. Until such data are available, it is important to recognize that – although being genuine – certain protein associations may fail to replicate or show opposite effects when compared across these two technologically different platforms. Thus, while orthogonal replication is confirmatory, lack of replication across datasets using these two different platforms does not invalidate the observed signals.

Fourth, we were in the unique position to probe for an overlap of prediagnostic biomarker profiles of PD and two other neurodegenerative diseases (AD and ALS) owing to the design of the EPIC4ND study. About one-third of PD biomarkers also showed consistent associations with incident AD and/or ALS, including RSPO2, SPAST, and PYDC1. Even though effect sizes correlated only modestly across diseases, the enrichment of shared biomarkers - especially between PD and AD - suggests partially overlapping prediagnostic mechanisms. While a limited biomarker overlap has recently also been reported by the GNPC for the symptomatic phase of these two diseases^25,26^; our results are the first to demonstrate that this is already detectable in the prediagnostic phase. Notwithstanding these overlaps, we emphasize that most identifed biomarkers were specific to PD, and the PD-specific protein prediction score was only predictive for PD but not for AD as demonstrated in the independent AGES cohort. Fifth, it appears that disease-related mechanisms are already active and detectable in blood more than two decades before diagnosis. A wide array of *in silico* analyses supporting the biological plausibility of many of our biomarker findings showed, for instance, an overrepresentation of key PD-relevant processes, including vesicle trafficking, dopaminergic neuron biology, and immune signaling. Furthermore, cell-type enrichment analyses implicated the involvement of microglia and peripheral immune cells already in the earliest disease phases, aligning with current neuroinflammation models of PD^27,28^. Moreover, our PPI analyses suggest that the prediagnostic biomarkers interacted more frequently than expected by chance with known PD proteins and correlated with *SNCA* expression, strongly emphasizing their functional relevance to the disease. Along these lines, several of our newly detected prediagnostic biomarkers had been implicated in other studies in PD before. For example, TCL1A^29^, GPC4^30^, LCN2^31^, AMN^32^, CXCL11^33^, CUL4B^34^, FCAR^35^, BECN1^36^, and NAMPT^37^ have previously been reported as potential blood biomarkers in (prevalent) PD in studies with smaller sample sizes. In addition, some of the prediagnostic PD biomarkers were reported to show differential expression in post-mortem brain tissue from PD patients compared to controls (e.g., LCN2^38^, GJA1^39^, TIMP1^40^, AMN^32^). Lastly, several of our top biomarker proteins are implicated in microtubule dynamics (TPPP2), vesicle trafficking in the presynapsis (VAMP3), mitochondrial functions (ACAD8), dopaminergic neuronal function and maturation (GPC4, FGF8) and immune functioning/neuroinflammation (CXCL12, IL11, TIM1, and TCL1A), which are all highly relevant in PD pathophysiology^41^. A detailed summary of key experimental findings and supporting evidence, can be found in the **Supplementary Material**.

Sixth, our druggability assessments revealed that nearly half of the FDR-significant biomarkers show evidence for small-molecule interactions. While blood-based biomarkers of brain diseases (like PD) do not automatically translate into therapeutic targets, this assessment may help prioritize candidates for future functional studies and repurposing efforts. The list of biomarkers we identified as correlating between blood and CSF may also aid in prioritization efforts.

Lastly, using machine-learning, we developed a PD-specific protein risk score comprising eight proteins, which showed robust predictive value in external validation within the AGES cohort within a follow-up of up to 17 years. Along these lines, existing tools, such as the updated MDS criteria (specialist settings) and the PREDICT-PD algorithm (broader screening)^42,43^ based on combinations of lifestyle, genetic, and/or clinical variables show promise but remain insufficient, particularly for detecting prediagnostic PD more than five years prior to diagnosis (e.g., ref. ^44^). These tools are continuously evolving, and integrating our PD-specific protein risk score offers an immediate translational next step to improve the classification of prodromal PD.

Our study has several strengths, including representing the largest collection of incident PD proteomic datasets to date, an exceedingly long-term follow-up, rigorous case ascertainment, the inclusion of several external datasets ensuring population and platform cross-validations, as well as extension to other disease phenotypes and diseases. Notwithstanding, the results of our study must be interpreted with the following potential limitations in mind. First, use of distinct proteomic platforms with different technologies, particularly SomaScan versus Olink, and coverages, comparatively limited case numbers despite large replication cohorts, usage of different blood sample types (citrate plasma, EDTA plasma, or serum) as well as differences in study design (e.g., recruitment, case ascertainment, follow-up times) may have contributed to non-replication of some biomarkers. At the same time, the biomarkers that did show replication across the datasets analyzed can be considered particularly robust. Second, while the SomaScan 7K assay offers extensive coverage, it is semi-quantitative. This means that signal changes may reflect different proteoforms rather than absolute protein levels. Mass spectrometry-based approaches are needed to clarify the underlying biology. Third, although we carefully adjusted for key covariates in numerous sensitivity analyses, residual confounding remains possible. Fourth, lacking neuropathological confirmation, some PD cases may have been misclassified - a common problem in large cohorts and a general limitation in epidemiological studies. However, given that medical charts of PD cases were reviewed by clinical specialists, the risk of misclassification in EPIC4PD is likely lower than in cohorts such as UKB. Finally, all datasets consisted exclusively or predominantly of non-Hispanic White participants. While the ARIC study also included a subset of Black participants (n=2,625) and their exclusion did not affect effect estimates, the sample size was too limited to allow for dedicated ethnicity-specific analyses of this subset. Thus, while our findings are likely generalizable to populations of European ancestry, their applicability to other ethnic groups requires further assessment.

In conclusion, our study newly identifies and validates a number of robust blood-based biomarkers that predict PD several decades before clinical onset. Our results underscore the importance of applying prospective cohort designs with incident case ascertainment to detect genuinely *pre*diagnostic biomarkers. By capturing early molecular changes, our work lays the foundation for developing novel prevention strategies applying precision medicine to PD and potentially other neurodegenerative diseases.

## Data Availability

Data can be accessed by external researchers after approval by the EPIC4ND working group and the EPIC steering committee. Potential collaborators can contact the EPIC4ND working group chair, Christina Lill and EPIC administrator Sherry Morris.

## Acknowledgements

The authors thank all EPIC participants and the countless scientists who have contributed to the EPIC cohort over the past 30 years. We thank the National Institute for Public Health and the Environment (RIVM), Bilthoven, the Netherlands, for contributing cases and ongoing support to the EPIC study. We sincerely thank Bertrand Hemon for his assistance with data management, and Prof. Paolo Vineis, Prof. Beate Ritz, Prof. Martin Wolkewitz, Prof. Klaus Berger, Prof. Valentina Gallo, Dr. Mark Frasier, and Dr. Lazaros Belbasis for helpful discussions.

## Funding

Updates of the PD case ascertainments, generation and processing of proteomic data were funded by the Michael J Fox Foundation (#008994 to C.M.L. and E.R.). Part of the proteomic data were also generated with funding from the Cure Alzheimer’s Fund (to C.M.L. and L.B.), and the ‘CReATe-Clinical Research in ALS and Related Disorders for Therapeutic Development’ Consortium (to C.M.L. and L.B.). CReATe (U54 NS092091) is part of the Rare Diseases Clinical Research Network (RDCRN), an initiative of the Office of Rare Diseases Research (ORDR), NCATS and funded through collaborations between NCATS, NINDS, and the ALS Association. Intramural funding was provided by the Interdisciplinary Centre for Clinical Research, University Münster (to C.M.L.). Additional funding was supplied by the Interdisciplinary Centre for Clinical Research, University Münster (to C.M.L., Lil3/001/25). C.M. Lill was supported by the Heisenberg program of the DFG (DFG; LI 2654/4-1).

The coordination of EPIC-Europe is financially supported by International Agency for Research on Cancer (IARC) and also by the Department of Epidemiology and Biostatistics, School of Public Health, Imperial College London which has additional infrastructure support provided by the NIHR Imperial Biomedical Research Centre (BRC). The national cohorts are supported by Associazione Italiana per la Ricerca sul Cancro-AIRC-Italy, Italian Ministry of Health, Italian Ministry of University and Research (MUR), Compagnia di San Paolo (Italy); Dutch Ministry of Public Health, Welfare and Sports (VWS), the Netherlands Organisation for Health Research and Development (ZonMW), World Cancer Research Fund (WCRF), (The Netherlands); Instituto de Salud Carlos III (ISCIII), Regional Governments of Andalucía, Asturias, Basque Country, Murcia and Navarra, and the Catalan Institute of Oncology - ICO (Spain); Cancer Research UK (C864/A14136 to EPIC-Norfolk; C8221/A29017 to EPIC-Oxford), Medical Research Council (MR/N003284/1, MC-UU_12015/1 and MC_UU_00006/1 to EPIC-Norfolk; MR/Y013662/1 to EPIC-Oxford) (United Kingdom). Previous support has come from “Europe against Cancer” Programme of the European Commission (DG SANCO). SomaScan® data were generated under Master Research Agreement, 14th December 2021, between Imperial College London and SomaLogic Inc. SomaLogic were not involved in analyzing or interpreting the data; or in writing or submitting the manuscript for publication.

National Institute on Aging (NIA) contracts N01-AG-12100 and HHSN271201200022C (for Vi. G.) and Althingi (the Icelandic Parliament) financed the AGES-Reykjavik study. IHA and Novartis have collaborated on proteomics research since 2012. This study was also funded by the NIA (1R01AG065596-01A1 to Vi. G., 2R01AG065596-03 to Va.G.), the University of Iceland Postdoctoral Fund (to E.A.F.) and the Icelandic Research Fund (2511008-051 to Va.G.). This research was supported, in part, by the Intramural Research Program (IRP) of the NIA. K.A.W. and F.L. are supported by the NIA IRP. ARIC is carried out as a collaborative study supported by National Heart, Lung, and Blood Institute (NHLBI) contracts (75N92022D00001, 75N92022D00002, 75N92022D00003, 75N92022D00004, 75N92022D00005). The ARIC Neurocognitive Study was additionally supported by U01HL096812, U01HL096814, U01HL096899, U01HL096902, and U01HL096917 from the NIH (NHLBI, NIA, National Institute of Neurological Disorders and Stroke [NINDS], and National Institute on Deafness and Other Communication Disorders [NIDCD]).

### Disclaimer

Where authors are identified as personnel of the International Agency for Research on Cancer / World Health Organization, the authors alone are responsible for the views expressed in this article and they do not necessarily represent the decisions, policy or views of the International Agency for Research on Cancer / World Health Organization.

## Authors’ contributions

Study concept and supervision: LB, PF, MJG, ER, CML; acquisition/contribution of data: SM, JMH, MG, AJ-Z, MJS, CT-S, NC-C, AV, DP, SS, TK, NW, RK, RT, RV, SP, GM, CS, NF, GNPC, VK, ViG, HG, LM, LMW, IT, VaG, KAW, PF, ER, MJG, CML, data analysis and interpretation: JH, AS, SM, EAF, FL, VV, RM, RK-L, VD, OO, LD, FH, YZ, FA, KSB, PMK, JMH, NW,TH, SB, AE, MH, VaG, LB, OR, LMW, IT, ViG, KAW, PF, ER, MJG, CML; drafting the manuscript: JH, SM, CML. Critical revision of the manuscript for content: all co-authors

## METHODS

### Discovery cohort and study design

The EPIC cohort is a large multi-center prospective study that recruited 519,978 participants (70.5% women, mostly aged 35-70 years at baseline) from 23 centers across ten European countries. With approximately 30 years of follow-up, EPIC was originally established to explore the relationships between diet and cancer, but it has since expanded to study other chronic conditions, including neurodegenerative diseases^12,13^. Briefly, baseline data including information on diet, physical activity, alcohol and tobacco use, past and current illnesses, reproductive history, and anthropometric measurements were collected between 1991 and 2000. Citrate plasma samples from ∼75% of all participants were obtained using standardized protocols, then aliquoted and stored long-term in liquid nitrogen (–196 °C) at a central biobank housed at the International Agency for Research on Cancer (IARC)/World Health Organization in Lyon, France. Since baseline recruitment, participants have been followed up regularly by data linkage and/or questionnaires at each center. The EPIC study was approved by the ethical committee of IARC and by the ethical review boards of each participating study center. All participants signed a written informed consent.

The ‘EPIC for Parkinson’s disease’ case-cohort (EPIC4PD) analyzed in this study is embedded within EPIC and aims to prospectively identify molecular biomarkers in PD^11^. The case ascertainment protocol has been described previously^11,50^. It was based on a source population of 157,288 subjects from ten EPIC centers across five European countries including Spain (Murcia, Navarra, and San Sebastian), Italy (Florence, Turin, and Varese), Netherlands (Utrecht and Bilthoven), UK (Norfolk/Cambridge), and Germany (Heidelberg). Potential cases were ascertained based on data linkage through a combination of center-specific methods using hospital records, general practitioner records, mortality records, drug repositories, and questionnaire data followed by re-review of medical records by expert neurologists for PD. Since the initial ascertainment, 163 incident PD cases were added following an update of the case ascertainment in selected centers (Heidelberg, Florence, Varese, Turin, Utrecht, Bilthoven)^11^. The subcohort for EPIC4PD was randomly drawn from these centers from EPIC-interact, which is a random subcohort within EPIC^51^. The data freeze for the current project was set to the time point of diagnosis of the last patient per center. For the purpose of identifying predictive PD biomarkers, we applied a washout phase after blood draw of two years prior to diagnosis to avoid reverse causation resulting in an exclusion of 23 PD patients.

### Generation and quality control of proteomic data in EPIC4PD

Protein abundances were measured in citrate plasma samples using the SomaScan 7K (v4.1) platform (SomaLogic, Inc.). SomaScan is a high-throughput proteomic platform that uses modified nucleotides (Slow Off-rate Modified Aptamers - SOMAmers) which specifically bind to protein targets and quantifies them in relative fluorescence units (RFUs) using DNA microarray^52^. The 7K version quantifies 7,596 aptamers targeting 6,432 unique proteins^53^. Measurements of overall 17,841 EPIC participants were performed at SomaLogic (Boulder, CO, USA) as previously described. Participants included also individuals who were selected for other endpoints (such as cancer). All samples were processed in a single procedure. SomaScan assays were performed on 96-well plates, with 11 control wells per plate used to monitor batch effects, accuracy, precision, and background. Each plate included five pooled calibrator replicates, three quality control (QC) replicates, and three buffer replicates. In addition, 233 replicates of one EPIC QC sample were included. Readout was conducted using Agilent hybridization and scanning technology. To control readout variability, twelve hybridization control aptamers were added during elution. Initial QC was conducted by SomaLogic and included normalization of protein RFUs based on the following steps: hybridization normalization, intraplate median normalization, plate scaling and calibration, and adaptive normalization to a population reference.

We computed intraclass correlation coefficients (ICCs) for all 7,596 aptamers based on the 233 EPIC replicate measurements. Aptamers were removed if they were not a protein (n=46) [Hybridization Control Elution (n=12), non-biotin (n=10), non-cleavable (n=4), spuriomer (n=20)] or non-human (n=261). Aptamers were further excluded if both (i) the ICC was below 0.5 and (ii) if the aptamer levels were below the limit of detection (LOD) in more than 90% of the samples (n=4). If the measured value of an aptamer was below the LOD, it was set to LOD/2. The final number of quality-controlled aptamers was 7,285 (targeting 6,381 unique proteins). Samples for which any SomaLogic normalization scale factor was outside [0.4-2.5] (n=247) and those detected as 0s using an approach based on PCA and the local outlier factor statistic (https://privefl.github.io/blog/detecting-outlier-samples-in-pca/) were excluded (n=0 for the method excluding outliers and n=5 for capping outliers). Plate correction was further applied through a residual approach: for each SOMAmer, its measurements were corrected for plate effect estimated in linear mixed effect models adjusted for center, age, sex, BMI, smoking status, and incidence of multiple cancer types, CVD, T2D, neurodegenerative diseases and death, to preserve possible biological variation due to these factors. Finally, only the samples belonging to the EPIC4ND case-cohort were retained (n=6,543 for both outlier handling methods).

Measurements of each SOMAmer were log_10_-transformed and further centered and scaled so that their mean was 0 and their standard deviation was 1 in the subcohort. Measurements deviating more than 5 SD from the mean of the normalized log_10_-transformed data were either excluded or ‘capped’ i.e., set to the value of 5 SD from the normalized log_10_-transformed mean. The first approach is based on the assumption that outliers represent technical or biological variation unrelated to PD (e.g., outliers due to blood type or sex or genetic variation not associated with PD) obscuring any association signal nearer to the mean. However, as we also acknowledge that outliers may represent strong PD biomarker signals, in the discovery phase, we equally ran all statistical analyses by capping these outliers. For all statistically significant findings, log_10_-transformed values were visually inspected (**Supplementary Figure 11**) to verify the appropriateness of the data transformation and the cutoff for the outlier removal. In the subsequent text and main tables, protein names based on results using capped data are labeled with an ‘cap’.

### Statistical analysis in EPIC4PD

The final effective EPIC4PD case-cohort included 4,538 EPIC4PD participants including 574 incident PD cases with 7,285 aptamers (corresponding to 6,381 proteins) available for statistical analysis. We used Cox proportional hazard regression analyses to examine the association of 7,285 aptamers with instantaneous risk of PD using age as time scale and Prentice weights to take into account the case-cohort setting ^54^. Our basic model was stratified (i.e., separate baseline hazard functions estimated) for 5-year age intervals, sex, and center. Cox proportional hazards regression was performed using three distinct time windows relative to disease onset: the full follow-up time period (2-28 years from baseline to diagnosis/censoring), as well as two shorter periods therein: 2-10 and 10-20 years. Furthermore, we performed sex-specific analyses after stratification for men and women using the full follow-up time period. Differences between sex-stratified effect estimates were tested by interaction analyses as previously described^55,56^.

Proteomic results were defined as significant at a FDR of 0.05. Furthermore, we defined “suggestively significant biomarker signals” as those findings that did not withstand FDR control in the proteomic analyses but had a p-value <10^−3^ and demonstrated consistency across all EPIC4PD analyses - specifically, at least two nominally significant (p<0.05) results and all effect estimates pointing in the same direction of effect.

In sensitivity analyses for all FDR-significant results, we tested a ‘full’ adjustment model including the baseline covariates school education (no education/primary school vs higher education), smoking status (dummy-coded as ‘never’, ‘former’, ‘current’), body mass index (BMI; weight/height^2^ [kg/m^2^]), and physical activity (dummy-coded as ‘inactive’, ‘moderately inactive’, ‘moderately active/active’). Furthermore, for FDR-significant results derived from analyses stratified for women, we adjusted for postmenopausal status (dummy-coded as premenopausal, postmenopausal, perimenopausal, and surgical postmenopausal status) and use of hormones for menopause (yes/no).

Time-varying effects of PD-associated proteins were explored using different approaches across the EPIC4PD dataset with 2-30 year follow-up. To visualize time-varying effects of top results in different time windows, HR pseudotrajectories were computed using 5-year sliding windows in one-year steps. For the analyses of the sliding windows close to age at diagnosis, the 23 participants diagnosed with PD within two years of baseline and previously excluded from the main analyses were re-included. Due to more limited numbers of cases toward the end of follow-up, the final window covered diagnoses occurring 16 to 25 years after baseline. Curves were fitted using the geom_smooth() function from the ggplot2 package in R.

Furthermore, exploratory analyses of time-varying effects of ALS-associated proteins showing FDR-significant association in either the proximal or distal time windows (see above) were performed by analyzing Cox models (Prentice-weighted, stratified for age, sex, and center) with a ‘protein level × log time-to-event’ interaction term, and and by fitting Cox models incorporating natural splines to assess non-linear time-dependent effects. For all these analyses, we used time since blood collection instead of age as the time scale. For the splines, we split the follow-up time into fine intervals (using the survSplit() function) and applied a time-varying coefficient (TVC) approach for each protein using spline interactions (protein × spline(time-to-event)). The Prentice-weighted Cox models were used adjusting for age (as a time-varying variable) and stratifying by sex and center. The nsk() function from the R package splines was used with 2 degrees of freedom. We evaluated the statistical evidence for time-varying effects using Wald tests.

### Independent replication of predictive biomarkers in prospective cohorts

Replication analyses of FDR-significant predictive biomarkers were performed on 64,856 individuals (1,034 incident PD cases) across three different prospective cohorts (AGES^57^, ARIC^58^, UKB^59^) based on Olink (UKB) and SomaScan (AGES, ARIC) data, respectively.

The first replication cohort, the Age Gene/Environment Susceptibility-Reykjavik (AGES-Reykjavik) Study^60^, is a population-based study of adults aged 65 years and older in Iceland, designed to investigate genetic susceptibility and gene-environment interactions related to age-associated diseases. Between 2002 and 2006, 5,764 participants, which were survivors of the original Reykjavik Study, underwent comprehensive interviews, examinations, and measurements. The study was approved by the Icelandic National Bioethics Committee (VSN: 00–063), the Icelandic Data Protection Authority, and the NIA IRB; all participants provided written informed consent. Proteomic profiling of baseline serum samples from the AGES cohort has been described previously^61^. After quality control and exclusion of prevalent PD cases, 5,308 participants had available proteomic data, further reduced to 5,090 when applying a 2-year washout period. Over the follow-up, 121 participants developed incident PD. In AGES, aptamer levels were log10-transformed. Further details on recruitment can be found in ref.^62^, and prevalent PD case ascertainment in ref.^63^. Medical and mortality records (ICD10: G20) up to 2019-03-19 were reviewed to ascertain incident PD cases.

The 2nd replication cohort, ARIC^58^, is an ongoing community-based cohort study, for which participants were enrolled from four communities across the United States: Washington County, MD; Forsyth County, NC; northwestern suburbs of Minneapolis, MN; and Jackson, MS. The ARIC participants who had SomaScan 5K data available (4,953 aptamers passing quality control) comprised 11,205 middle-aged participants at baseline of whom 164 developed incident PD applying a washout period of two years and had SomaScan data in EDTA plasma samples available. For details on the cohort recruitment and ascertainment of incident PD cases therein please see ref.^64^ and the **Supplementary Material**. All participants provided appropriate written consent.

The 3rd replication cohort, UKB^59^, is a general population cohort study, which performed proteomic profiling of blood plasma samples using the Olink Explore 3072 platform (n=2,923 proteins, cohort size analyzed in this study: n=48,561 UKB participants including 749 incident PD cases). The plasma samples were collected in volunteers aged 40 to 69 years at their first attendance to a UKB assessment center in 22 centres across the United Kingdom between 2006 to 2010. Follow-up began from the date of attendance to an assessment center (field 53) to the first recorded date of diagnosis (ICD-10 code G20) in the patient-linked hospital episode statistics (fields 41270 and 41280) or death registration (fields 40001 and 40002), whichever occurred first, with the last recorded date in January 2023. Due to small case numbers in other strata, the analysis was restricted to white participants (field 21000). Participants with missing proteomics data were removed prior to each model. Due to smaller sample sizes in the restricted time windows, we did not analyze top EPIC4PD biomarkers in AGES for the time-window of 2-10 years of follow-up (n cases=26) and for ARIC for the time window of 10-20 years of follow-up (n cases=32).

Consistent with the EPIC4PD analyses, outliers in AGES and UKB were either capped at or excluded beyond 5 SD, depending on the approach used for the specific biomarker to be replicated. In AGES and UKB, outliers were either capped at or excluded beyond 5 SD, in line with EPIC4PD analyses. Outliers in ARIC were capped at 5 SD. Cox proportional hazards regression models were harmonized for AGES, ARIC and UKB with EPIC4PD: The Cox model was based on age as time scale and stratified (separate baseline hazard functions estimated) for age in 5-year categories, race (in ARIC), and sex. Significance for the replication arm was defined at α=0.05. As sensitivity analysis, we also performed the same analyses only on white participants in ARIC (excluding 2,625 black participants). However, as results did not change substantially, we only report results on all participants in this study.

We explicitely note that the smaller amount of replication data of distal biomarkers (i.e. falling into the 10-20-year pre-diagnosis window) is a a result of shorter follow-up periods in some replication cohorts, reducing the number of available replication datasets (excluding ARIC, and, in many instances, UKB), as well as limited case numbers (AGES) after time-window stratification. Specifically, while all time-window independent biomarkers were assessed in 3-4 independent datasets, for 14 out of 19 biomarkers from the 10-20 year window, only 1-2 replication datasets were available. Consequently, we have intentionally exercised caution and avoided highlighting confirmatory (but limited) external replication evidence to prevent overinterpretation.

### Indirect replication and characterization of prediagnostic EPIC4PD biomarkers for clinical phenotypes in GNPC

The FDR-significant prediagnostic biomarkers identified in EPIC4PD across the three follow-up periods and after stratification for sex were assessed for an association with prevalent PD leveraging a large case-control dataset of the GNPC (335 prevalent cases and 2,257 controls from 3 contributors) by re-analyzing the data described in the companion manuscript of Imam et al.^14^ All individuals included had undergone SomaScan 7K proteomic profiling as part of the GNPC initiative. For this analysis, we restricted inclusion to datasets that contained both PD cases and controls. Control participants were those without a documented clinical diagnosis of mild cognitive impairment, dementia, AD, frontotemporal dementia, PD, or ALS. To minimize bias from population stratification and ensure comparability with EPIC4PD, we only included white participants in datasets where the variable “race” was available. If no race information was provided, all participants were included. The protein data was log10-transformed and standardized and values above or below five standard deviations from the mean were excluded. Then, the case-control datasets were residualized using the removeBatchEffect function from the R package limma to regress out the effects of age, sex, and contributor, thereby controlling for confounding sources of variation in the high-dimensional data. A principal component (PC) analysis was then performed on the residuals (i.e., after having removed the effects of age, sex, dummy-coded contributor); based on the scree plot, the respective first two PCs were retained as additional covariates. Finally, logistic regression analyses were conducted on the original data, adjusting for age, sex, dummy-coded contributor, and the first two PCs (all results from the re-analyses with p<0.05 can be found in **Supplementary Table 24**). Analogous to the EPIC4PD analyses, prediagnostic biomarkers that were FDR-significant in one sex stratum were analyzed for the GNPC data within that specific sex group.

Concordance of effect estimates in EPIC4PD and GNPC (for all 47 significant prediagnostic biomarkers as well as for nominally significant biomarkers shared by both studies) we applied a one-sided binomial exact test. All analyses were performed in R.

We analyzed serum proteomic data from the Tracking Parkinson’s disease cohort using the SomaScan v4.1 (7K) assay (SomaLogic). Our cohort included 794 PD patients with serum samples and clinical progression data across the three study visits (at 0 months, 18 months, 36 months). To correct for batch effects introduced by collection site, we used the ComBat function from the sva R package (v3.50.0), with site as the batch variable, followed by log_10_-transformation. For association analyses with PD progression, we investigated associations between protein levels and two clinical outcomes: the Unified Parkinson’s Disease Rating Scale Part III (UPDRS-III) as a measure of motor function, and the Montreal Cognitive Assessment (MoCA) as a measure of cognitive function.

Cross-sectional associations were assessed using generalized linear models, adjusting for age, sex, and Levodopa Equivalent Daily Dose (LEDD) as a measure of treatment exposure. For longitudinal analyses across up to to three visits per participant, we applied linear mixed-effects models using the lmer() function from the lme4 R package. These models included a random intercept for participant ID to account for repeated measures, and fixed effects for protein level (log_10_ RFU), visit, age, sex, and LEDD. In additional sensitivity analyses, we included BMI and disease duration (time since diagnosis at baseline) as covariates. Protein associations were evaluated separately for each outcome, and p-values were adjusted for multiple testing using an FDR=0.05.

### Comparison to previous studies

We compiled a list of 291 unique candidate biomarkers previously linked to PD based on findings from studies that employed a range of different methodologies (several proteins were nominated more than once): These included i) 38 proteins from the only previous proteomic study on future PD performed on 2,920 proteins based on Olink measurements in the UKB^10^, ii) 10 of ∼2,700 analyzed blood plasma proteins (7 derived from Olink and 3 from SomaScan studies) recently proposed as potential predictive biomarkers for PD by a comprehensive MR study^16^, iii) 18 proteins nominated by a recent MR and colocalization study^65^, iv) 109 proteins corresponding to nominated functional candidate and/or the nearest genes located in 78 genetic risk loci described in the most recent Caucasian genome-wide association study for PD ^17^, and v) 127 preselected neurodegenerative disease candidate biomarkers included in the ‘CNS NULISA panel’ (Alamar, Inc.). Proteins available in our SomaScan 7K data from this list (n=180) were tested for association in EPIC4PD. Results were controlled applying an FDR=0.05 across all 180 candidate proteins available for testing in our SomaScan 7K data.

### Cross-disease comparisons

For cross-disease comparisons between PD and AD as well as between PD and ALS, we performed Cox proportional hazard regression analyses for all FDR-significant PD proteins in EPIC4AD (1,926 EPIC participants including 652 incident AD cases during a follow-up of 2-29 years) and EPIC4ALS (4,715 EPIC participants including 187 incident ALS cases during a follow-up of 2-30 years) using Prentice-weighted Cox proportional hazard regression analysis stratified for 5-year age groups, sex, and center, as described above. Results were controlled at an FDR=0.05 in the context of all tested top biomarkers per disease. Furthermore, we assessed whether significant associations in incident AD and ALS were enriched among all proteins that were at least nominally significant in PD (based on the full follow-up period, outliers excluded), using a one-sided Fisher’s exact test. To assess whether overlapping nominally significant proteins exhibited concordant effect directions more often than expected by chance, we performed a one-sided binomial test. Furthermore, we assessed the correlation of effect sizes between traits by calculating Pearson’s correlation coefficients based on the beta estimates. To prevent bias in the correlation due to the discontinuity introduced by limiting the analysis to nominally significant results, we aligned effect directions by mirroring the beta coefficients: for proteins with negative betas in PD, we multiplied all corresponding betas in EPIC4PD and GNPC by –1 (generating ‘mirrowed beta values’).

### Brain expression data and protein abundance of top biomarkers

Gene expression and protein abundance of top PD biomarkers across tissues and in the brain were assessed based on RNA-Seq and immunohistochemisty (IHC) data obtained from the Human Protein Atlas (HPA v24.0, based on Ensembl version 109, accessed May 6th, 2025 at https://www.proteinatlas.org/about/download) ^66^. FDR-significant proteins were probed for tissue expression using bulk RNA sequencing (RNA-Seq) data across 50 human tissues available as gene-level ‘Consensus RNA data’ (rna_tissue_consensus.tsv.zip). Furthermore, brain expression was assessed based on gene-level bulk RNA-Seq data across 13 brain regions based on samples from the Human Brain Tissue Bank (Semmelweis University, Budapest; rna_brain_region_hpa.tsv.zip) and based on single nucleus RNA-Seq (snRNA-Seq) data across 11 brain regions (rna_single_nuclei_cluster_type.tsv.zip). Protein measurements for the FDR-significant biomarkers across 45 human tissues were obtained from HPA based on IHC using tissue microarrays (normal_ihc_data.tsv.zip). To visualize expression patterns of the top biomarkers, heatmaps were generated using hierarchical clustering, with expression values scaled per gene highlighting tissue-specific variation.

### Gene set, cell type enrichment and network analyses

Gene set enrichment analyses were performed based on the GO classification system using the PANTHER resource (Protein ANalysis THrough Evolutionary Relationships; version 19.0; released 2024-06-20; https://geneontology.org/). A list of all unique proteins that showed a suggestive signal in EPIC4PD and was uniquely mapped to a gene by PANTHER (n=154) was tested for enrichment for ‘biological process’, ‘molecular function’, and ‘cellular component’ GO categories as well as ‘Reactome pathways’. The statistical overrepresentation test was conducted using Fisher’s exact test with the reference list set to all analyzed SomaScan 7K proteins uniquely mapped by PANTHER (n=6,332). To avoid spurious associations, only GO categories with at least ten proteins present in the background list were considered.

Cell enrichment analyses were performed using the the Expression Weighted Cell Type Enrichment (EWCE) method to predict the likely primary disease-associated cells based on our prioritised proteins^67^. This approach generates a probability distribution of gene expression for specific cell types by comparing a target gene list to a background gene set. We applied EWCE to our protein lists and to either single-cell mouse brain expression data^68^ or RNA expression data from 23 selected cell types sourced from the Human Protein Atlas (dataset: rna_single_cell_type.tsv.zip)^66^. Each analysis was controlled at an FDR=0.05. CorrelationAnalyzeR was used to investigate the coexpression of all FDR-significant prediagnostic biomarkers. Coexpression with *SNCA* was specifically examined in 12,712 normal GEO samples from immune cells and 3,198 normal GEO samples from brain.

### Protein-protein interaction network analysis

Furthermore, protein-protein interaction (PPI) networks were built using the STRING database (v12.0, accessed May 6th, 2025 at https://string-db.org)69 based on all FDR-significant EPIC4PD biomarkers and plot using igraph (v2.1.4). Interactions with a minimum interaction score of 0.4 were based on the following sources: ‘textmining’, ‘experiments’, ‘databases’, ‘co-expression”, “neighborhood’, ‘gene fusion’, and ‘co-occurrence’. Furthermore, to examine the list of suggestive prediagnostic proteins in relation to known PD genes, we utilized 109 proteins described in the most recent genome-wide association study for PD (see above)^17^. P values were calculated based on the observed interactions compared to 10,000 random permutations. Nodes not connected to the largest component were removed from the PPI network Figure 3.

### Drug Repurposing

We leveraged the Open Targets platform^19^ to explore druggability and drug-repurposing potential for all FDR-significant, putative prediagnostic PD biomarkers. Open Targets integrates multiple data sources to systematically link genes and proteins with drug molecules and disease associations. We queried the database focusing on small-molecule evidence, excluding other therapeutic modalities such as biologics or gene therapies. Specifically, approximately 17,000 small-molecule drugs and 21,087 protein targets were screened for evidence of interaction with the 47 FDR-significant biomarkers. Evidence of druggability was defined by reported direct binding or predicted high-quality pharmacological modulation supported by curated experimental, 3D protein structure or clinical data within the Open Targets resource.

### Prediction

To assess the proteins’ performance in risk stratification for PD, we performed variable selection in a Cox proportional hazards model using the Least Absolute Shrinkage and Selection Operator (LASSO) for variable selection and regularization. Specifically, EPIC4PD (excluding the Heidelberg center) served as the training set. Incident PD cases were Prentice⍰weighted to account for the case⍰cohort sampling and class imbalance, and penalized Cox models were fitted using LASSO as implemented in the R package glmnet with cv.glmnet(family = “cox”, type.measure = “C”). The amount of penalization (λ) was tuned via 5⍰fold cross⍰validation using the C⍰index as the performance criterion. For model selection, each candidate λ from the cross⍰validation grid was evaluated in a Cox model restricted to participants from the Heidelberg center, and the λ yielding the highest C⍰index in Heidelberg defined the final set of proteins and their regression coefficients. These coefficients were used to compute the PD proteomic risk score (PPRS), which was then evaluated in the independent external AGES cohort in Cox models adjusted for age and sex. To this end, the improvement of the C-index from the baseline model (comprising age and sex only) to the baseline model + the top PPRS was quantified. This approach controls for age effects and avoids selecting aging markers as PD predictors. To assess whether the PPRS captures general neurodegenerative process, we also quantified the ΔC-index for AD in AGES with and without inclusion of *APOE* epsilon 4 genotypes.

